# Michigan Neural Distinctiveness (MiND) study II protocol: Investigating age-related neural dedifferentiation longitudinally and in Alzheimer’s pathology

**DOI:** 10.1101/2025.11.10.25339905

**Authors:** Esther Kim, Molly Simmonite, Kayla Wyatt, Quan Zhou, Mark Zuppichini, Noah Reardon, Adriene M. Beltz, Benjamin M. Hampstead, Stephan F. Taylor, Thad Polk

## Abstract

**Background:** A growing body of research has found that neural representations of task stimuli are less distinctive in older adults relative to younger adults, a phenomenon known as age-related neural dedifferentiation. The original funding period of the Michigan Neural Distinctiveness (MiND) study aimed to investigate the scope, causes, and consequences of neural dedifferentiation. We recruited a sample of healthy older adults (aged 65+ years) and younger adults (aged 18-29) and administered fMRI (to measure neural distinctiveness), MRS (to measure GABA), and substantial behavioral testing (to measure cognitive, motor, and sensory functions). We found that neural distinctiveness was lower in older vs. younger adults, that reduced GABA was associated with this neural dedifferentiation, and that behavioral performance was associated with both neural dedifferentiation and reduced GABA. In this current paper, we describe the rationale and methods for the second phase of the MiND study, in which we: 1) investigate age-related trajectories of change in neural distinctiveness, GABA, and behavior longitudinally; and 2) explore how neural differentiation is related to Alzheimer’s disease pathology.

**Methods:** In the longitudinal aim, we are re-contacting participants from the original MiND funding period. We aim to test participants at two additional time points approximately 3-5 years apart using the same assessments (fMRI, MRS, and behavioral tests). We plan to have approximately 150 participants with two time points and 100 participants with three time points. We will also recruit new participants who will be tested twice, approximately 3-5 years apart in order to further increase our sample size and power. We predict to have approximately 250 new participants with one time point. We will then test whether neural distinctiveness, GABA, and behavioral performance decline longitudinally with age.

To explore neural dedifferentiation in Alzheimer’s disease pathology, we plan to recruit 100 MCI participants who are already undergoing PET imaging to determine amyloid beta and tau burden. These patients are then completing our fMRI, MRS and behavioral testing sessions so that we can examine associations between Alzheimer’s pathology and our measures of neural distinctiveness, GABA, and behavior.

**Discussion:** This line of research has the potential to lead to new insights into how aging affects the mind. In particular, it could shed light on the role that neural dedifferentiation, GABA, and Alzheimer’s disease pathology play in age-related behavioral declines.

**Trial Registration:** This study was registered with the ISRCTN registry on November 11, 2024. The registration number is ISRCTN16528440.

## Background

The world is facing an aging crisis. In 2010, 13% of the United State’s population was over the age of 65, by 2040 it is expected that this percentage will increase to more than 20% [1]. Healthy aging is associated with significant behavioral declines, even in the absence of significant disease. Working memory, long-term memory, speed of processing, problem solving and perception have all been reported to exhibit age-related decline from age 20 onward [2, 3]. Aging is also the single greatest risk factor for Alzheimer’s disease. Thus, as the older population grows, so too will the number of cases of this disease, and the large burden of dealing with it. Alzheimer’s disease is characterized by the accumulation of amyloid beta plaques and neurofibrillary tangles; however, it is well established that the behavioral symptoms of Alzheimer’s disease typically appear many years after the plaques and tangles start affecting the brain, often decades later [4]. A central challenge is to understand the neural mechanisms underlying behavioral impairments in both healthy aging and Alzheimer’s disease. Developing such an understanding is the first step toward designing interventions to address the significant societal burdens associated with these impairments.

Previous research has suggested that age-related declines in neural distinctiveness may be an important factor underlying behavioral impairments associated with aging. Using fMRI, several studies have demonstrated that the neural activity patterns in the ventral visual cortex elicited by faces, places, and words were significantly more distinctive in young adults in comparison with healthy older adults [5–8]. This phenomenon is referred to as age-related neural dedifferentiation. In older adults, individual differences in fMRI-based neural distinctiveness account for up to 30% of the variance in behavioral performance on fluid processing tasks [6]. In addition, single-neuron recordings from the primary visual cortex in non-human primates have shown that, while the activity in response to different orientations and directions was selective in young macaques, the older macaques responded much more non-selectively, showing strong responses to all stimuli [9]. Interestingly, following the application of either the inhibitory neurotransmitter gamma-aminobutyric acid (GABA) or the GABA agonist muscimol, the same cells in the old macaques showed strong selectivity for stimulus orientation [10]. Conversely, the neurons in the young monkeys that were strongly orientation selective remained so following the application of GABA, but selectivity was abolished following the application of the GABA antagonist bicuculline, which made their firing patterns resemble those of the neurons of the old monkeys at baseline. These findings demonstrate that changes in GABA activity can cause changes in neural selectivity.

Given these findings, the original funding period of Michigan Neural Distinctiveness (MiND; [11]) study aimed to explore the scope, causes, and consequences of age-related changes in neural distinctiveness. While most previous studies had focused on ventral visual cortex, we used functional magnetic resonance imaging (fMRI) to not only replicate that finding [12], but also to demonstrate reduced neural distinctiveness in older adults in auditory [13] and motor cortices [14]. Additionally, there were significant positive cross-region correlations between neural distinctiveness measures in different brain regions, indicating not only that neural dedifferentiation extends beyond visual cortex, but also that these changes appear in tandem across the brain [15].

Using magnetic resonance spectroscopy (MRS) to estimate GABA levels, the original MiND study also revealed that GABA levels in the visual, auditory, and somatosensory cortices are reduced in older adults relative to young adults. Furthermore, individual differences in GABA levels in specific brain regions were related to individual differences in neural distinctiveness in that same brain region. For example, GABA levels in ventral visual cortex were positively related to the distinctiveness of fMRI patterns in response to faces and houses in the same region [12]. Similarly, auditory cortex GABA levels were positively correlated with the distinctiveness of neural patterns elicited by music and foreign speech [13]. Our findings are consistent with the hypothesis that age-related declines in GABA are associated with age-related declines in neural distinctiveness.

Here, we outline the next phase of the MiND study, which seeks to address limitations of the original study, and to extend the project to examine GABA-based neural dedifferentiation within the context of Alzheimer’s disease pathology. Our first aim is to explore longitudinal trajectories of both neural distinctiveness and GABA concentrations in older adults. Longitudinal aging studies have many advantages over more common, cross-sectional studies as they capture the trajectory of change over time within individuals rather than just the differences between age groups. They are therefore not confounded by cohort effects, that is, differences between groups that are unrelated to age (e.g. childhood experiences, educational experiences, nutrition, etc.). While these advantages are widely recognized by aging researchers, they are typically uncommon as they take many years to conduct. By inviting the older participants of the original funding period of the MiND study back to repeat the fMRI, MRS and behavioral assessments two more times, we will examine longitudinal trajectories of change in GABA and neural distinctiveness in the visual, auditory, and motor cortices, as well as longitudinal changes in behavior on a battery of cognitive tasks.

Our second aim is to investigate the role of GABA-based neural dedifferentiation in the context of Alzheimer’s pathology. The original MiND study investigated GABA and neural distinctiveness exclusively in the context of healthy aging, but some evidence suggests that they may also play a role in Alzheimer’s disease and related dementias. For example, Maass et al. [16] used fMRI to estimate neural activity while participants completed a memory task during which they indicated whether images of scenes and objects were identical to, or slightly different from, previously presented pictures. The same participants also underwent PET imaging to estimate amyloid and tau burdens. Activation in a posterior-medial network involving spatial memory was significantly less selective to scenes in tau-positive older adults than in tau-negative older adults or young adults, demonstrating neural dedifferentiation in Alzheimer’s pathology. Furthermore, individual differences in neural distinctiveness were associated with memory performance; reduced neural distinctiveness was associated with reduced memory performance.

There is also evidence that GABA may play an important role in Alzheimer’s disease and MCI. In particular, several post-mortem studies [17–25], studies of cerebrospinal fluid [26–28], and MRS studies have reported reduced GABA levels in a variety of brain regions in patients with presumed Alzheimer’s disease or MCI [29, 30].

Inspired by these findings, we will measure neural distinctiveness, GABA, and behavioral performance in patients with MCI from whom amyloid and tau PET images have already been acquired. We will then test whether neural distinctiveness and GABA are significantly lower in MCI patients than in age-matched controls and whether neural distinctiveness mediates the relationship between neuropathology and behavior. We will add the scene-object discrimination task used by Maass et al [16] into our fMRI task battery alongside the visual, auditory, and motor tasks included in the original MiND protocol, as well as acquiring GABA estimates from a posterior medial MRS voxel.

In summary, the second phase of the Michigan Neural Distinctiveness (MiND) study will investigate age-related trajectories of change in GABA, neural distinctiveness, and behavior as well as directional relationships between these factors, via longitudinal measures. We will also examine neural differentiation, GABA and behavior in the context of Alzheimer’s pathology. Table 1 summarizes the main hypotheses being investigated.

**Table 1.**
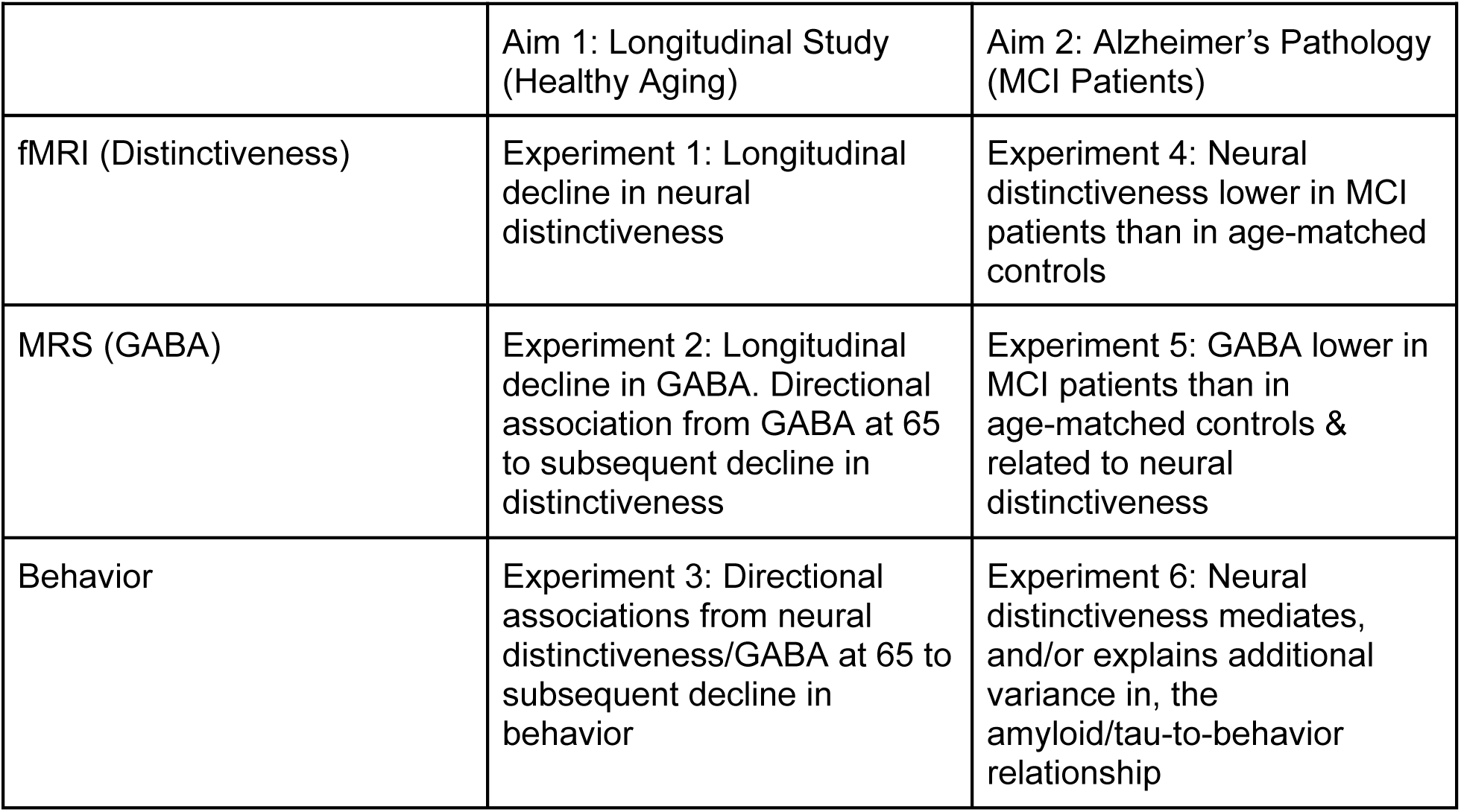
Project Hypotheses.

## Methods

### Participants

All participants are older adults aged 65+ years old, right-handed, native English speakers. Sessions are conducted in University of Michigan’s Functional MRI Laboratory at the Bonisteel Interdisciplinary Research Building, Ann Arbor Lakes Building 1, and the Ann and Robert H. Lurie Biomedical Engineering Building in Ann Arbor, Michigan.

For the longitudinal arm of the study, participants who took part in the original MiND study [11] are being re-contacted, and invited to repeat the study sessions. Participants who took part in the original MiND study (Wave 1) are invited to take part in a second set of sessions approximately 3-5 years later (Wave 2), and a third set of sessions another 3-5 years after that (Wave 3). In addition, new Wave 1 participants are recruited from Ann Arbor and the surrounding area, via flyers posted throughout the community (libraries, college campus, community centers, etc.). The University of Michigan also hosts a website (https://umhealthresearch.org/) where individuals can learn about research studies at the university that are currently recruiting participants, and submit their contact details.

For the Alzheimer’s pathology arm of the study, older adults with a research diagnosis of Mild Cognitive Impairment (MCI) are referred to the MiND study from collaborating studies being conducted at the University of Michigan, including the NIA P30 funded Michigan Alzheimer’s Disease Research Center. Following referral to the MiND study, potential participants are contacted via phone by study coordinators and screened.

Full inclusion and exclusion criteria are listed in **Table 2**. Since MCI participants are difficult to recruit, and since we have limited pools from which to recruit Wave 2 and Wave 3 participants (i.e, the participants that have completed Wave 1), exclusion criteria are relaxed somewhat (refer to **Table 2**), to ensure we are able to recruit sufficient samples.

**Table 2.** Inclusion & Exclusion Criteria.

|  | Cognitively Normal, Wave 1 Participants | Mild Cognitive Impairment participants, and Wave 2 and 3 participants |
| --- | --- | --- |
| Inclusion criteria | • 65+ years old, right-handed, native English speaker<br>• Normal (or corrected-to-normal) vision, hearing, and motor control<br>• MRI scan compatible | • 65+ years old<br>• MRI scan compatible |
| Exclusion criteria | • Use of hearing aids, color blindness, motor control problems, psychotropic medication, use of estrogen therapy,<br>• Current depression/anxiety or occurrence within the past 5 years,<br>• Concussion with unconsciousness for 5 min or more.<br>• More than 7 drinks per week<br>• History of drug or alcohol abuse or addiction<br>• Weight greater than 250 pounds<br>• MRI incompatibility (claustrophobia, foreign metallic objects, pacemaker etc)<br>• Familiar with languages in auditory task (Sinhalese, Macedonian, Marathi/Hindi, Persian, Ukrainian, Swahili). | • Weight greater than 250 pounds<br>• MRI incompatibility (claustrophobia, foreign metallic objects, pacemaker etc) |

### Power calculations

To examine longitudinal change in neural distinctiveness and GABA, we assume at least 2 times points per participant, a correlation of at least 0.37 in within-person measures (reflecting within-person stability), residual variance of 0.5 across all assessments, and a slope estimate of |0.3| (reflecting a difference from 0) in a two sided mixed model with a Bonferroni-adjusted alpha of .0167 for multiple comparisons of three neural distinctiveness indicators (visual, auditory, and motor; since we will not have longitudinal data for our memory task) [31]. These assumptions are based on small to medium effect sizes. Under these assumptions, the proposed sample size of 150 results has 88% power to detect significant change over time, according to a Monte Carlo simulation with 1000 replications. This is consistent with independent simulations that suggest sample sizes of 100-200 are sufficient to detect moderate effects in similar models [32].

Furthermore, using cross-sectional correlations between GABA and neural distinctiveness and between distinctiveness and behavior from our own work [6, 12, 13] as an estimate of effect size, 150 participants will provide 99% power to detect links between GABA intercepts and neural distinctiveness slopes and between GABA/distinctiveness intercepts and behavior slopes with a Bonferoni-adjusted alpha of .0167 [33, 34]. Sensitivity analyses adjusting for gray matter volume and adding sex and its interaction with the neural predictors also have high power (96%). Power remains acceptable (>80%) for correlations as small as .25. These calculations are based on only 2 time points, but note we aim to have 3 timepoints in approximately 110-120 of the participants, further increasing power.

With a sample of 100 MCI participants and 150 controls, we will have at least 80% power to detect medium-sized (*d* = .42) differences in neural distinctiveness and GABA between the groups in general linear models with covariates of gray matter volume and age [33, 34], even with a Bonferoni-adjusted Type I error of .0125 for multiple comparisons (visual, auditory, motor and memory). This detectable effect size is smaller than previously reported effects (*d* = 0.77) [30]. This sample size also provides power to detect small-to-medium sized relationships (*r* = .21) between GABA and neural distinctiveness and a small-to-medium sized incremental prediction of cognitive function from neural distinctiveness above and beyond amyloid and tau.

### Study Design

Following recruitment, individuals are screened via telephone interview and online surveys, including the Cognitive Failures Questionnaire [35] to determine their eligibility to participate in the study. Once determined eligible, participants are scheduled to complete three in-person testing sessions, each on separate days. All three sessions are typically done within a month; Session 1 and Session 2 may be consecutive days but Session 2 and Session 3 are typically conducted 5-7 days apart. This is done so that the participants’ Session 2 fMRI data may be acquired and preprocessed to be used to prescribe voxel locations for their Session 3 MRS scan. Prior to the first session, MiND study coordinators read the consent form to participants, and answer any questions that participants may have via phone call. To ensure informed consent, MiND study coordinators ensure participants have understood the consent document by asking questions to probe understanding. Should participants be unable to demonstrate understanding of the informed consent document, their participation in the study is discontinued.

Session 1 lasts for ∼120 minutes and consists of consenting, followed by cognitive and behavioral testing. At the beginning of the session, participants are asked to read through the consent form, ask any final questions regarding the details of their participation and complete the consent form. Following this, participants who have not received a research diagnosis, or who require an updated research diagnosis, are administered the National Alzheimer’s Coordinating Center Uniform Data Set Version 3 (NACC UDS3) battery [36] by trained research staff. Each research diagnosis uses the National Alzheimer’s Coordinating Center (NACC) criteria and is considered valid for 9 months. The rest of the session is spent completing tasks from the MiND cognitive and behavioral battery. For all participants, Session 2 consists of 30 mins of behavioral testing, 15 mins of preparation for the MRI scan (instruction, practice, etc.) and a 60 min MRI session. In Session 3, all participants complete a 90 min MRS scan session. Those participants who require a research diagnosis and complete the NACC UDS3 battery during Session 1, also spend a further 60 mins during Session 3 completing any outstanding items from the MiND cognitive and behavioral battery.

### Research Diagnosis Assessments

All participants taking part in the MiND study are tested using the NACC UDS3 battery [36], which is appropriate because many are co-enrolled in the National Institute on Aging (NIA) P30 funded Michigan Alzheimer’s Disease Research Center (MADRC) longitudinal cohort. These data are used to determine their research diagnosis of either cognitively normal (CN), MCI or dementia of the Alzheimer’s type (DAT). If a participant meets the criteria for DAT, then they are excluded from the study. To ascertain each participant’s research diagnosis, the NACC UDS3 battery is administered by trained research staff. The results of this testing, alongside general health information, are reviewed by a collaborating Board-Certified Clinical Neuropsychologist (BMH). The testing comprises of the following measures:

#### 1. The Geriatric Depression Scale (GDS)

The GDS assesses depression in older adults. It typically takes five minutes to administer and consists of 15 questions that prompt a “Yes” or “No” response from the participant. Each question gauges the presence of depressive symptoms experienced in the past week, including the day it is administered. Five questions reveal depressive symptoms when answered “No” and the remaining questions do so when answered “Yes”. If the response reveals a sign of depression it is given 1 point. A total score ranging from 0 to 4 is considered normal, while a score between 5 and 15 indicates some level of depression (mild to severe). If a participant’s score exceeds 5, or question 11 is answered “No”, a risk assessment must be completed by a certified clinician. The risk assessment involves further questioning to determine the participant’s ability to continue testing.

#### 2. Montreal Cognitive Assessment (MoCA) [37]

The MoCA is a test of cognitive function using 13 brief tasks, in total taking about 10 minutes. Attention and concentration are tested with a letter sequence task where participants tap their hand when they hear “A” and a number subtraction task. Executive function is assessed through the alternating trail-making task, a verbal phonemic fluency task, and an abstraction task involving explaining word pairs’ similarities. Memory is evaluated by recalling a list of five nouns immediately and again after a delay of five minutes. Language skills are measured with a naming task (identifying animals) and a complex sentence repetition task. Visuoconstructional skills are tested by drawing a three-dimensional cube and a clock. Lastly, orientation is assessed by asking the participant to report the current time and place. The MoCA is scored out of 30 points, with a score of 26 or higher considered in the normal range.

#### 3. Craft Story 21 Recall (Immediate and Delayed)

This test assesses episodic and semantic memory. The examiner reads a short story containing 25 items of information. The participant is asked to retell the story from memory immediately and again following an 18-22 minute delay period. Participants receive credit for each informational bit recalled verbatim and for accurate paraphrases, with a maximum score of 25 for the immediate retelling and 25 for the delayed retelling.

#### 4. Benson Complex Figure Test (Copy, Recall, Recognition)

This test evaluates visual spatial cognition in assessing visuoconstructional and visual memory functions. A paper with a figure is presented to the participant. The examiner provides a pen and instructs the participant to copy the design. When the design is completed, the participant is told that they must remember this design and are given approximately 5 seconds to review the design they copied. The participant must then draw the design from memory 10-15 minutes later. Subsequently, the participant is shown four figures and must identify the figure initially copied. This test is scored based on each element’s accuracy and placement in the drawing.

#### 5. Number Span Test (Forward and Backward)

This is a working memory test. The participant is read a series of numbers at one-second intervals and is asked to repeat the series in the same order and reverse order. The sequences presented range from 2 to 9 numbers. Testing is discontinued when two consecutive trials of the same number length are incorrect.

#### 6. Category Fluency

This test measures semantic memory, verbal fluency and language. The participant is asked to name as many examples as possible within a given semantic category (animals and vegetables) in 60 seconds. Scoring is based on the number of valid responses provided. For animals, credit is not received for repetitions and mythical animals. For vegetables, credit is not received for repetitions, prepared vegetable products and spices.

#### 7. Trail Making Test (Part A and Part B)

Participants are instructed to connect a sequence of consecutive targets as quickly and accurately as possible. For Part A, participants are given a sheet with 23 numbered circles. They are given a pencil without an eraser and instructed to draw a line from one number to the next, in order. For Part B, participants are given a sheet with 23 circles containing numbers or letters. They must take the pencil without an eraser and draw a line from one circle to the next alternating in order between number and letters. For both tests, participants are also instructed to do their best to not lift the pencil as they move from one circle to the next. The test is timed with performance measured by the time taken to complete each task. Part A is discontinued after 150 seconds and Part B is discontinued after 300 seconds.

#### 8. Multilingual Naming Test (MINT)

This test measures the ability to name visual objects. Participants are presented with 32 images of commonplace items. These items are presented as black-and-white line drawings. The stimuli are presented beginning with relatively easier items and progress to more challenging ones. After each image is presented, the participant has 20 seconds to give a spontaneous response before a semantic cue is given. If the participant still can’t identify the item after the semantic cue is given, a phonemic cue is provided which includes the sound of the first letter of the item’s name. The task is discontinued after 6 consecutive failures, which occur if the participant can’t name the item or requires a phonemic cue 6 times in a row. The task score is calculated by adding the number of spontaneous correct responses and correct responses following a semantic cue.

#### 9. Verbal Fluency: Phonemic Test

For the verbal fluency task the participant is given a letter of the alphabet and is instructed to name as many words as they possibly can that start with that letter in the allotted 60 seconds. This is done for the letters C, F, and L. Credit is not given for repetitions, non-words, grammatical variants, proper nouns, or numbers. Scores are calculated as the sum of words named for each letter and are adjusted for age and education.

### MiND Cognitive & Behavioral Assessment Battery

Participants perform the following battery of tasks to assess cognitive, motor and sensory abilities. Tasks completed during each session and task order are listed below in **Table 3**. All NIH Toolbox tasks are administered using the NIH Toolbox for Assessment of Neurological and Behavioral Function iPad App [38]. Further information about the NIH Toolbox scoring methodology can be found on the NIH toolbox website’s scoring guide. Additionally, **Table 4 provides brief** descriptions of each task and indicates the behavioral domain and subdomain that each task is intended to assess.

**Table 3.**
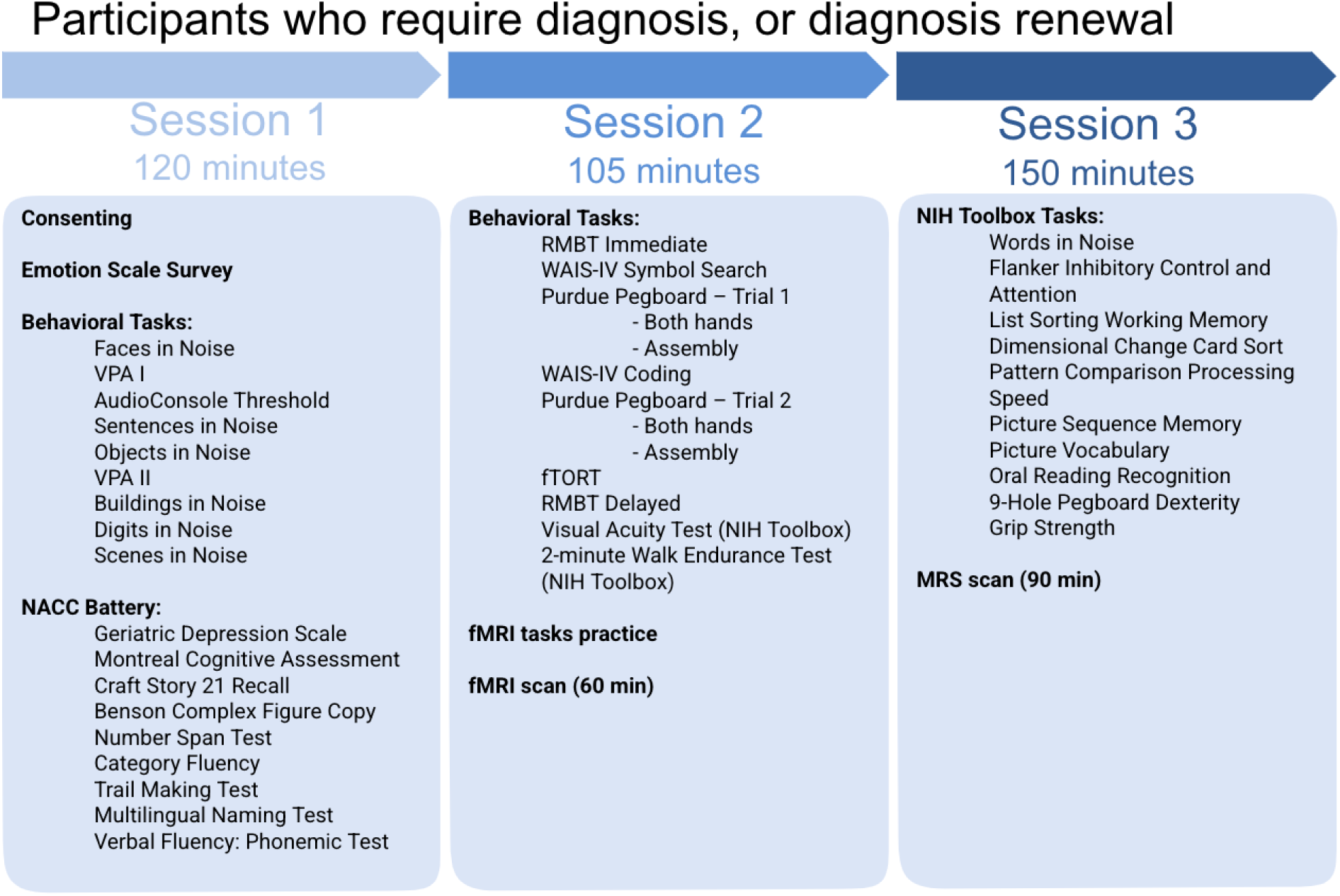
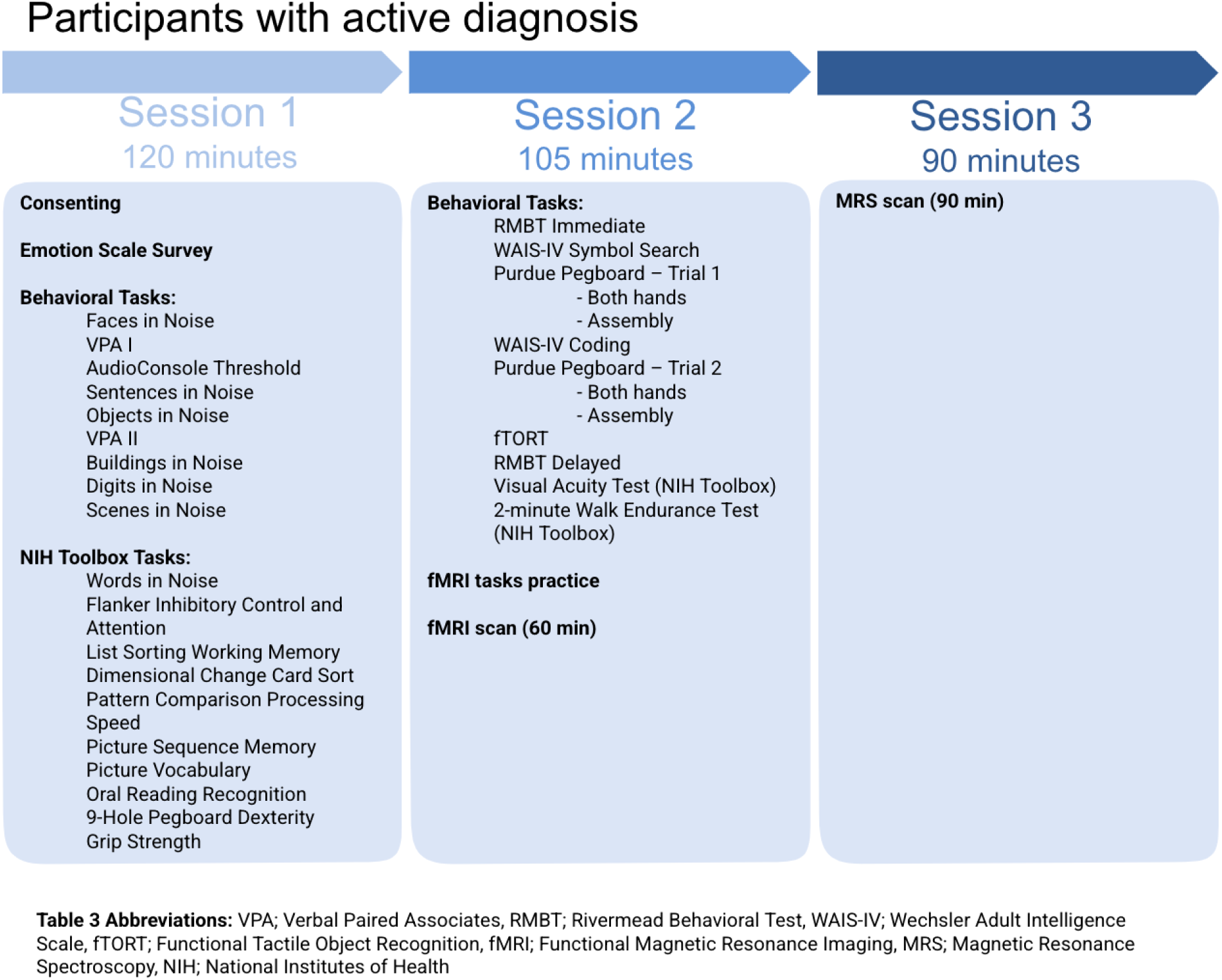
Session & Task Order.

**Table 4.**
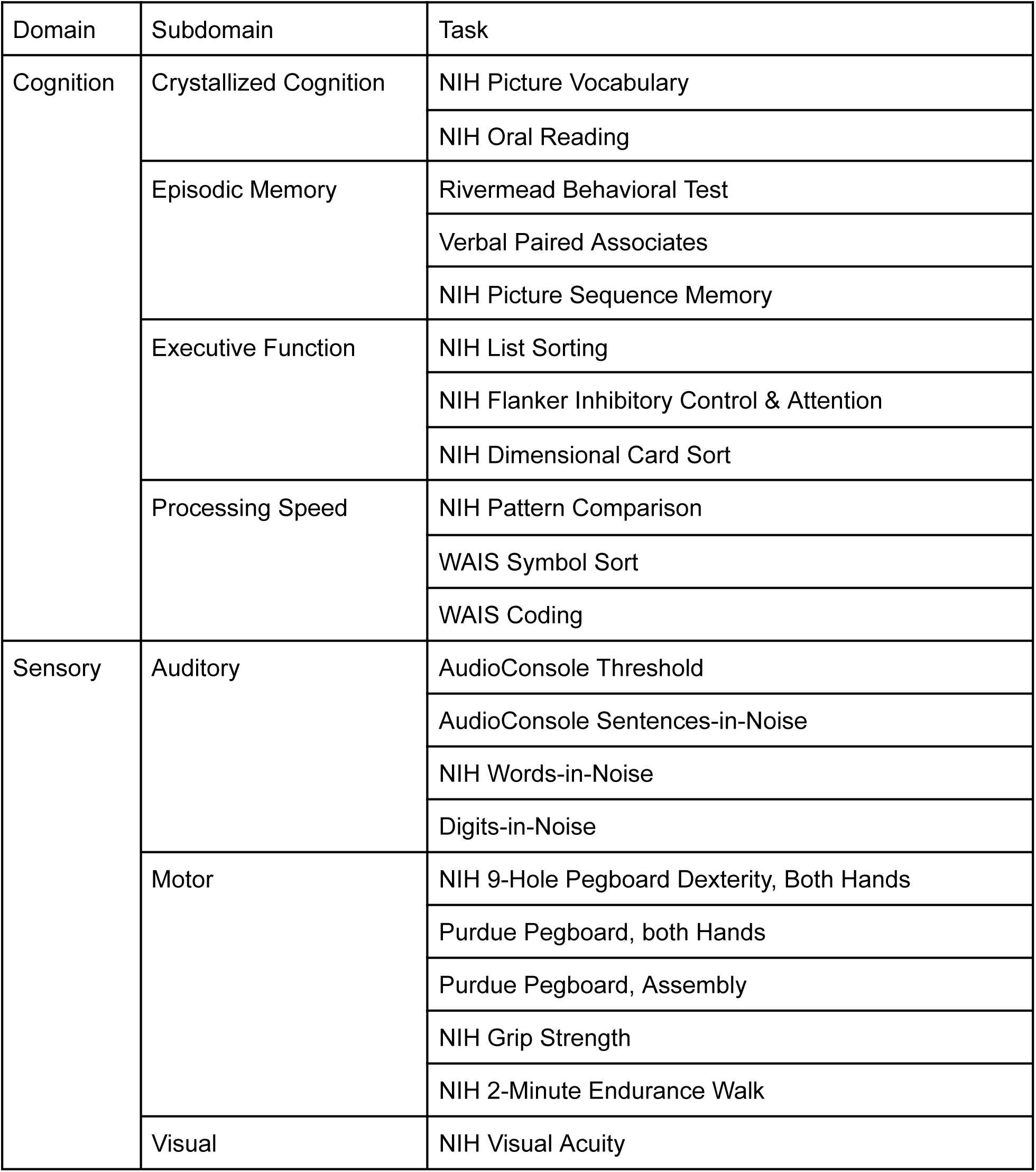
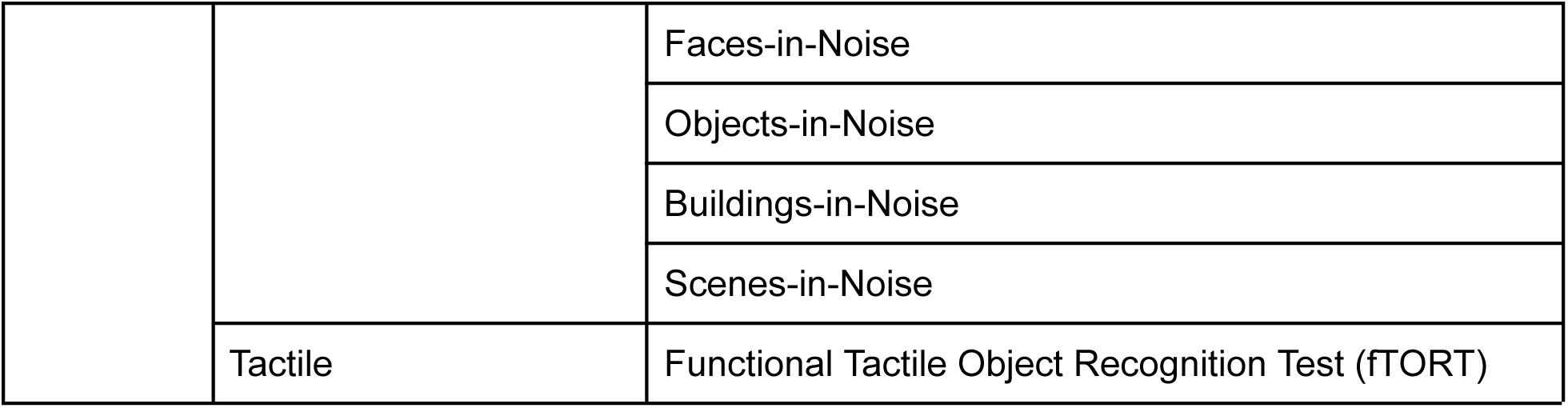
Behavioral Tasks.

| Domain | Subdomain | Task |
| --- | --- | --- |
| Cognition | Crystallized Cognition | NIH Picture Vocabulary |
|  |  | NIH Oral Reading |
|  | Episodic Memory | Rivermead Behavioral Test |
|  |  | Verbal Paired Associates |
|  |  | NIH Picture Sequence Memory |
|  | Executive Function | NIH List Sorting |
|  |  | NIH Flanker Inhibitory Control & Attention |
|  |  | NIH Dimensional Card Sort |
|  | Processing Speed | NIH Pattern Comparison |
|  |  | WAIS Symbol Sort |
|  |  | WAIS Coding |
| Sensory | Auditory | AudioConsole Threshold |
|  |  | AudioConsole Sentences-in-Noise |
|  |  | NIH Words-in-Noise |
|  |  | Digits-in-Noise |
|  | Motor | NIH 9-Hole Pegboard Dexterity, Both Hands |
|  |  | Purdue Pegboard, both Hands |
|  |  | Purdue Pegboard, Assembly |
|  |  | NIH Grip Strength |
|  |  | NIH 2-Minute Endurance Walk |
|  | Visual | NIH Visual Acuity |
|  |  | Faces-in-Noise |
|  |  | Objects-in-Noise |
|  |  | Buildings-in-Noise |
|  |  | Scenes-in-Noise |
|  | Tactile | Functional Tactile Object Recognition Test (fTORT) |

### Visual function

#### 1. NIH Visual Acuity

This test measures binocular visual acuity. Participants are instructed to wear corrective lenses or glasses, if required. They sit 3 meters away from an iPad screen and letters appear on the screen one at a time. Participants verbally state the letter they see and the size of the letter changes depending on the accuracy of their answer. This task establishes each participant’s visual ability baseline.

#### 2. Vision-in-Noise Tests

Four vision-in-noise tasks are administered on a Dell laptop, using the Psychophysics Toolbox [39] in MATLAB[40]. Details of the stimuli for each vision-in-noise task are listed below. Each task comprises 50 trials, in which a fixation cross is presented for 500ms, followed by a black and white stimulus presented in dynamic Gaussian noise for 500ms. For each task, the stimulus is a member of one of two categories, which must be indicated by the participant. Once the participant indicates a response, the next trial begins. There are 15 levels of Gaussian noise used, and each task begins on the 5th level. Using a staircase procedure, the level of noise is increased or decreased, depending on the accuracy of the participant’s responses. For each task, the participants’ score is the average noise level on the final 40 trials.

A. **Faces**. The stimulus picture is a male face on 50% of trials and a female face on 50% of trials. Participants are instructed to press “1” on the laptop keyboard with their left index finger if they think the picture is a male face or press “0” with their right index finger if they thought it was a female face.
B. **Objects**. The stimulus picture is an office item on 50% of trials and a food item on 50% of trials. Participants press “1” on the keyboard with their left index finger if they think the picture was an office item or press “0” with their right index finger if they think the picture is a food item.
C. **Buildings**. The stimulus picture is a house on 50% of trials and an apartment on 50% of trials. Participants press “1” on the keyboard with their left index finger if they think the picture was a house or press “0” if they think the picture was an apartment building.
D. **Scenes**. The stimulus picture is an urban scene on 50% of trials and a nature scene on 50% of trials. Participants press “1’on the keyboard with their left index finger if they think the picture was urban or press “0” with their right index finger if they think the picture was a nature scene.

### Auditory function

#### 1. NIH Words-in-Noise

Participants wear over-the-ear noise canceling headphones and hear single words presented with varying levels of background noise. Participants are asked to say the word they hear to the best of their ability, while the examiner types their responses on an iPad. Participants complete the task twice - once with words presented to their left ear and once with words presented to their right ear.

#### 2. AudioConsole Threshold

This task determines participants’ baseline hearing thresholds at different frequencies. Participants are asked to press a response button when they can hear a tone. This task is administered using a Dell laptop using the Oscilla USB-350SP PC-based audiometer software with TDH-39 Headphones, utilizing the Hughson Westlake Automatic Hearing Test.

#### 3. AudioConsole Sentences-in-Noise

Examiners use a Dell laptop using Oscilla USB-350SP PC-based audiometer software with TDH-39 headphones. Examiners work with the participant to set up the task at a comfortable volume, for each ear. The volume decibel is recorded. Then participants hear a sentence and are instructed to repeat the sentence to the best of their ability. The participant goes through three lists of sentences while the examiner types their responses on the laptop.

#### 4. Digits-in-Noise [41]

This task utilizes a Dell laptop, MATLAB and over-the-ear noise canceling headphones. The volume is set to a comfortable level by the participant and examiner prior to starting, and the volume is recorded by the examiner. The participant hears groups of 3 numbers in both ears with varying signal-to noise-ratios (SNR). The decibel level of the digits varies based on their previous answer accuracy and the level of noise is kept constant.

### Tactile function

#### 1. Functional Tactile Object Recognition Test (fTORT) [42]

Participants are asked to identify everyday objects through the sense of touch. They are given a poster with pictures of the items and a corresponding number. They are instructed to place their hands in a box, to obstruct their view of the target object, and to indicate the number of the object they are touching. Their answer and their response time is recorded.

### Motor function

#### 1. NIH 9-Hole Pegboard Dexterity, Left Hand & Right Hand

Participants are presented with a board with a pile of pegs and 9 holes. They are instructed to place one peg into each hole as quickly as possible, one at a time. After each hole has a peg, participants then take out each peg as quickly as possible, one at a time. Participants are timed and do this task for each hand.

#### 2. Purdue Pegboard, Both Hands (<u>Purdue Pegboard User Instructions</u>)

Participants are presented with the Purdue Pegboard. They are instructed to pick up a peg, one in each hand, at the same time. They place each peg, at the same time, in the first row of holes and continue to pick up and place pegs down the rows. Participants will do as many pegs as they can until they are told to stop; they have 30 seconds and the examiner will record the number of rows completed.

#### 3. Purdue Pegboard, Assembly (Purdue Pegboard User Instructions)

Using the same Purdue Pegboard, participants are given a different task. They are instructed to use both hands and create assemblies consisting of 3 items: peg, collar and washer. Participants are instructed to 1) pick up a peg with their right hand, 2) place the peg in the hole, and pick up a washer with their left hand, 3) place the washer on the peg and pick up a collar with their right hand, 4) place the collar on the same peg and pick up another washer with their left hand, 5) finally place the washer on the same peg, completing the assembly. While placing the final washer, the participant picks up a peg with their right hand to start the next assembly in the next hole. The examiner the number of assemblies and items placed.

#### 4. NIH Grip Strength

Participants are seated in a chair with their feet flat on the ground and have their arms at their sides, bent at a 90° angle. They squeeze a Jamar Plus Digital dynamometer as hard as they can for 3 seconds, for each hand.

#### 5. NIH 2-Minute Endurance Walk

In this task, two cones are placed 25 feet apart. Participants are instructed to stand at one cone and walk back and forth around the two cones, as quickly as they can without running or hurting themselves, for 2 minutes. The examiner records the distance walked by counting laps around the cones.

### Processing Speed

#### 1. NIH Pattern Comparison

Participants are shown two images and are instructed to determine whether the two images are the same, as quickly as possible. The response options are yes or no. Participants respond as many times as they can in 85 seconds.

#### 2. Wechsler Adult Intelligence Scale (WAIS) Symbol Search [43]

Participants are presented with rows consisting of 2 target symbols on the left and 5 response symbols on the right. They are instructed to draw a line through the response symbol that matches one of the target symbols. If there are no matching symbols, participants are instructed to draw a line through a “No” box. Participants have 2 minutes to complete as many rows as possible.

#### 3. Wechsler Adult Intelligence Scale (WAIS) Digit-Symbol Coding [43]

Participants are given a key consisting of a set of numbers-symbol pairs. They are also given rows of numbers with blank boxes below them. Participants are instructed to write in the symbol corresponding to each number using the key. They are given 2 minutes to complete as many as possible.

### Executive function and working memory

#### 1. NIH List Sorting

There are two phases of this task. In the first, pictures of food or of animals are presented on an iPad. Participants are instructed to sort the images based on how big they are in real life, from smallest to biggest. In the second phase, the images are a random sequence of both food and animals. Participants are instructed to again sort the images by size, but to sort the food first and then sort the animals.

#### 2. NIH Flanker Inhibitory Control and Attention

Participants are presented with a row of arrows each of which is pointing either left or right. They are instructed to indicate the direction that the middle arrow is pointing as quickly as possible, using a button on an iPad screen. Trials may be congruent, in which all arrows point in the same direction, or incongruent, in which the middle arrow (the target) points in the opposite direction to the surrounding arrows (the flankers). The task consists of 20 trials in total, 40% of which are incongruent.

#### 3. NIH Dimensional Change Card Sort

In this task, participants are presented with one target image and two response images. The two response images correspond to either the color or the shape of the target image. During the trial, the word COLOR or SHAPE will be shown on screen and then the participant is instructed to select the response image accordingly. There are 30 trials, 23% of which are color trials.

### Episodic memory

#### 1. Rivermead Behavioral Test (RMBT) [44]

There are two sections of this test: immediate recall and delayed recall. For the immediate recall section, participants listen to a brief story (5 sentences). They are then asked to tell the examiner as much of the story they can remember. The delayed recall section is a surprise memory test that occurs roughly 20 minutes after the immediate recall. Participants are then asked to tell the examiner as much of the story they can remember.

#### 2. Verbal Paired Associates (VPA) [43]

This task consists of two sections that are conducted approximately 25 minutes apart. There is a list of 14 word pairs; some word pairs are related (e.g. sock-shoe), and some are not (laugh-stand). In the first section of the task, the administrator will read each word pair, out-loud, to the participant. Then, the administrator will say the first word of each pair and ask the participant what the corresponding second word is. Participants will receive feedback for each answer; if the answer is incorrect, the participant will be reminded of the correct answer. This task will repeat three more times.

The second section is a surprise memory test. In this second section, the examiner will again say the first word of the pair and ask the participant for the corresponding second word. However, in the second round, the participant will receive no feedback on the accuracy of their answer. Once finished with the original list, the examiner will then read a list of 40 word pairs. The participant is instructed to indicate if the stated word pair was in the original list from the first section. Both words must be correct and matching to be from the initial list.

#### 3. NIH Picture Sequence Memory

A sequence of 15 images is displayed on an iPad screen. After viewing the sequence, the participant is shown all the images at once and is asked to arrange the images in the order in which they originally appeared. Once finished, a second round of images are presented. In this second trial, the sequence consists of 18 images: 15 from the first round and 3 new images. Participants are asked to arrange the 18 images in the presented sequence.

### Crystalized intelligence

#### 1. NIH Picture Vocabulary

This test is a measure of general vocabulary knowledge. Participants hear an audio recording of a word and four pictures are presented on an iPad screen. They select the picture that best matches the meaning of the word.

#### 2. NIH Oral Reading Recognition

This test measures reading ability. Participants are presented a word on an iPad screen and are instructed to read the word out-loud. The test administrator uses a pronunciation guide and scores the response’s accuracy.

### MRI session protocol

MRI data is collected using a 3T GE Discovery Magnetic Resonance System with an 8-channel head coil at the University of Michigan’s Functional MRI Laboratory. The fMRI session lasts approximately 60 minutes, during which task-based and resting-state functional data, a high resolution structural image and diffusion-weighted imaging (DWI) data are acquired. Before beginning the MRI session, participants are shown previews of each of the tasks and allowed to practice outside the scanner.

Data acquired during this session are as follows, in order:

(1) **3-Plane Localizer** The localizer is generated with a 8-channel, 2D Spin Echo pulse sequence, FOV = 320 x 320 mm, Slice thickness = 10 mm with zero spacing, acquisition time approximately 10 seconds.
(2) **T1-weighted overlay** The overlay is generated with a 8-channel, 2D FSE-XL pulse sequence. TR = 2314.3 ms, TE = 24.0 ms, TI = 960 ms, Flip angle = 111 degree, FOV = 220 x 220 mm, 43 oblique slices with slice thickness = 3.0 mm with zero spacing, acquisition time approximately 144 seconds.
(3) **fMRI data** The T2-weighted images for all four functional tasks and resting state are collected using a 8-channel, 2D Gradient Echo (GRE) reverse spiral pulse sequence, TR = 2000 ms, TE = 30 ms, flip angle = 90, FOV = 220 x 220 mm, 43 oblique slices with slice thickness = 3 mm no spacing. Heart rate is also collected using a pulse oximeter, respiration is collected with a plethysmography (respiration belt), and any required responses are collected using a Celeritas 5-button fiber optic response unit. All participants complete the auditory task first followed by the resting-state scan. This is to ensure that the fMRI compatible earbuds are working correctly, and in addition to ensure that all participants resting-state data is collected after the auditory task, in accordance with data collection in the original MiND study. Following the resting-state scan, the memory, visual, and motor tasks are completed, in a counterbalanced order.

A. **Auditory Task** Participants are instructed to press a button with their right index finger every time they hear the target trial: hearing a tone/beep. The auditory task scan duration is 6 minutes and consists of six 20-second blocks of foreign speech clips, six 20-second blocks of instrumental music clips, and twelve 10-second blocks of no sound. The foreign languages used in the foreign speech blocks are Creole, Macedonian, Marathi, Persian, Ukrainian, and Swahili. Participants are screened to ensure that they are unfamiliar with these languages. The speech and music blocks are pseudo-randomized and the block order is the same for all participants. A no-sound block always follows each speech and music block. A target trial has a beep interjected into the speech or music block. There are six target trials: three during speech blocks and three during music blocks. Participants view a fixation cross on the screen for the duration of the task. This task utilizes an Avotec Conformal Headset to play the auditory stimuli.
B. **Resting State** Participants are instructed to relax, keep their eyes open and focus on the fixation cross displayed on the screen for the duration of the scan. The scan lasts 8 minutes.
C. **Memory Task** Participants are presented with sequences of 4 consecutive objects or 4 consecutive scenes. They are instructed to press a button with their right index finger or right middle finger depending on whether the stimulus was presented earlier in the sequence or not. The task was derived and modified from [45]. This task scan length is 6 minutes and has two runs, so a total of 12 minutes. There are 36 total sequences, 18 object sequences and 18 scene sequences. Each stimulus is shown for 3 seconds and the stimuli are separated by a fixation point. Interstimulus intervals are jittered between 0.6 - 4.2 seconds while intervals between sequences average 2.43 seconds to stress the end of the sequence.
D. **Motor Task** Participants are instructed to press a button with their right thumb every time they see a right-facing arrow and to press a button with their left thumb every time they see a left-facing arrow. This task’s duration is 6 minutes and has six 20 second blocks of a left-pointing arrow, six 20 second blocks of a right-facing arrow and twelve 10 second blocks of a fixation cross. Every arrow block is followed by a fixation block and the block order is pseudorandom, but the same for all participants.
E. **Visual Task** Participants are presented with blocks of faces and blocks of buildings. They are instructed to press a button with their right index finger every time they see a female face (during the face blocks) or an apartment building (during the building blocks). The scan length is 6 minutes and has six 20-second blocks of images of faces, six 20 second blocks of images of buildings, and twelve 10 second blocks of a fixation cross. Every face and building block is followed by a fixation block and the block order is pseudorandom, but the same for all participants. There are six target trials, 3 during face blocks and 3 during building blocks.
(4) **High Resolution Structural Image** A high-resolution T1-weighted structural image is collected using a 8-channel, 3D gradient-echo SPGR, BRAVO pulse sequence, TR = 11.4 ms, TE = 5.1 ms, TI = 500 ms, flip angle = 15, FOV = 256 x 256 mm, 156 axial slices with 1 mm thickness with no spacing (1 x 1 x 1 mm voxels, 152 axial slices), acquisition time 5:27 minutes.
(5) **Diffusion Weighted Imaging data** Diffusion weighted imaging (DWI) multi-shell data is collected using a 32-channel head coil and a diffusion-weighted 2D Spin Echo Pulse Sequence, TR = 4100 ms, TE = minimum, FOV = 240 x 240 mm, 81 axial slices with thickness 1.7 mm, 0 slice spacing, 102 diffusion directions: 6@b value=0, 6@b=500, 15@b=1000, 15@b=2000, and 60@b=3000, acquisition time 7:15 minutes.

### MRS session protocol

MRS data is collected using the same MRI scanner and head coil (8-channel) as the MRI scan session. The MRS session lasts approximately 75 mins total. Sessions begin with a 3-Plane Localizer and high-resolution T1-weighted structural scan using the same parameters as those acquired during the MRI session. MRS data from seven voxels of interest (VOIs) are then acquired using a MEGA-PRESS sequence [46] using the following parameters: TR = 1800 ms; TE = 68 ms (TE1 = 15 ms, TE2 = 53 ms); 256 transients (128 ON interleaved with 128 OFF) of 4096 data points; spectral width = 5 kHz; frequency selective editing pulses (14 ms) applied at 1.9 ppm (ON) and 7.46 ppm (OFF); FOV = 240 × 240 mm; voxel size = 30 × 30 × 30 mm. Acquisition time for each voxel of interest (VOI) is 8.5 minutes.

MRS data is collected from left and right visual voxels, left and right auditory voxels, left and right somatosensory voxels and a midline memory voxel. For each participant, MRS voxels are placed to ensure maximal overlap with peak fMRI activation during the visual, auditory, motor and memory tasks completed during the MRI session. For the visual, auditory and motor tasks, a GLM is performed on each task, contrasting each of the two experimental conditions with the fixation block. Then, the two contrast maps and the high resolution structural image acquired during the MRI session are used to place the voxels such that they capture the areas of highest activation for both conditions, in each hemisphere. For example, contrast maps of houses vs fixation and faces vs fixation during the visual fMRI task are used to place voxels in the left and right ventrovisual cortices. Contrast maps of speech vs no-sound blocks and music vs no-sound blocks during the auditory fMRI task are used to place voxels in the left and right auditory cortices. Contrast maps of right hand motor movements vs fixation blocks are used to place a voxels in the left somatosensory cortex and contrast maps of left hand motor movements vs fixation blocks during the motor fMRI task are used to place a voxel in the right somatosensory cortex. In an effort to replicate Maass et al. [16], the memory voxel was placed with reference to each participant’s peak activation in the precuneus during the scene vs. object contrast.

### Preprocessing Pipelines

#### Structural MRI

Freesurfer’s recon-all function is used to construct and segment a cortical surface for each participant from their T1-weighted anatomical image [47].

#### Functional MRI

##### Task-based fMRI preprocessing

Freesurfer’s FSFAST is used for preprocessing and first-level analyses of fMRI data. Preprocessing includes motion correction and spatial smoothing using a Gaussian kernel with full width half maximum (FWHM) of 5mm.

##### Resting state fMRI preprocessing

Resting-state functional MRI data is preprocessed using the default pipeline from the CONN toolbox [48]; this includes realignment, slice-time correction, outlier detection, segmentation, normalization to MNI space, and spatial smoothing. To further reduce the influence of motion-related artifacts on functional connectivity, an additional censoring step is applied post-preprocessing, following the recommendations of Power et al., [49]. Post-preprocessing, we evaluate the average BOLD signal fluctuations (z-scores) and framewise displacement (FD) on a per-participant basis. Outliers are identified using thresholds of 3 standard deviations for BOLD signal change and 0.3 mm for FD [49].

Resting state data is denoised using a denoising pipeline in the CONN toolbox. This paradigm combines two general steps: linear regressions of potential confounding effects in the BOLD signal and temporal band-pass filters. Potential confounding factors affecting the estimated BOLD signal are independently identified and removed for each voxel, participant, and functional run/session. This is achieved through Ordinary Least Squares (OLS) regression, which projects each BOLD time series onto a subspace orthogonal to the identified confounding effects. CONN’s toolbox default denoising pipeline utilizes an anatomical component-based noise correction (aCompCor) method and accounts for noise components from white matter and cerebrospinal fluid, estimates motion parameters, and removes outliers and first-order linear session effects. Temporal frequencies outside the range of 0.008 - 0.09 Hz are removed from the BOLD signal, in order to focus on slow-frequency fluctuations and to reduce the impact of physiological noise, head motions and other artifacts. This range filtering is applied through a discrete cosine transform windowing operation to minimize edge effects and is performed after linear regressions to prevent any frequency misalignment in the nuisance regression process [50].

After preprocessing, ROI-to-ROI connectivity analysis is conducted using the HCP-ICA atlas. This is done through independent component analysis (ICA) on data from the Human Connectome Project. The effects of aging on connectivity, both cross-sectional and longitudinal, are examined through multilevel modeling.

#### DWI

Diffusion-weighted multi-shell images (DWI) are preprocessed using MRtrix [51]. The preprocessing steps include denoising, unringing, motion and distortion correction (eddy current distortions), and bias field correction. The data is upsampled to an isotropic voxel size of 1.3 mm using b-spline interpolation and is intensity normalized across participants based on the median b = 0 s/mm^2 [52].

Fiber orientation distributions (FODs) are computed using multi-tissue constrained spherical deconvolution [53]. All participant FOD images are registered to the FOD template. Using this a white matter template analysis fixel mask is created with a peak value of 0.06. Then each FOD track is segmented (angle 22.5, maxlen 250, minlen 10, power 1.0) and the whole brain tractography is performed on the FOD template generating 20 million streamlines. Spherical-deconvolution informed filtering of tractograms is also applied, producing an output of 2 million streamlines.

#### MRS

To quantify GABA, we utilize the Gannet 3.3.2 MATLAB toolbox [54] for each of the seven MRS voxels. The data is time domain frequency and phase corrected using spectral registration and filtered with a 3-Hz exponential line broadening and zero-filled by a factor of 16. Gannet uses a five-parameter Gaussian-Lorentzian model to fit GABA peaks between 2.19 and 3.55 ppm. Metabolite concentration values are scaled to water and expressed in institutional units (IU). GABA estimates are corrected for the fraction of voxel that is cerebrospinal fluid (CSF) and for different concentrations of GABA in gray and white matter [55].

### Analysis Pipeline

#### Calculating Neural Distinctiveness

Neural distinctiveness estimates are made using the task fMRI data. In order to calculate neural distinctiveness, we utilize multi-voxel pattern analysis (MVPA) in functionally defined ROI (regions of interest). First, neural activity is estimated using GLM (generalized linear model) in FSFAST. In each task, responses to the two experimental conditions are modeled using a block design, including separate regressors for each of the experimental blocks convolved with a canonical hemodynamic response function. Specifically for each task, auditory: speech and noise, visual: faces and houses, motor: left and right finger tapping, and memory: objects and scenes.

Then, using FreeSurfer’s Cortical Parcellation, bilateral anatomical masks for each task are created for each participant using their T1-weighted structural image. For the auditory task, the mask includes the superior temporal gyrus, transverse temporal gyrus, bank of the superior temporal sulcus, and supramarginal gyrus. For the visual task, the mask includes the fusiform gyrus and the parahippocampal gyrus. For the motor task, the mask includes the precentral gyrus, postcentral gyrus and supramarginal gyrus. For the memory task, the mask includes perirhinal cortex and parahippocampal cortex.

We then use in-house MATLAB code to create functional regions-of-interest (ROIs) for each task and each participant. First, the vertices within each participant’s anatomical mask are sorted based on activation during the first experimental condition vs. rest. Then, these vertices are sorted a second time based on activation during the second experimental condition vs. rest. Lastly, the functional ROI is defined by alternating between the two sorted lists; this is done by taking the most active vertex from the first experimental condition, then adding the most active vertex from the other experimental condition, and so on. This alternating system continues until a target functional ROI size is reached. This method is used to define functional ROIs of different sizes (50 vertices, 100 vertices, … 10,000 vertices). The activation estimates from the created functional ROI is used to calculate neural distinctiveness of multi-voxel representations.

Neural distinctiveness is estimated by comparing the similarity of activation patterns within the functional ROIs during the same stimulus conditions (e.g., within different face blocks, within different house blocks, etc.) to the similarity of activation patterns during different stimulus conditions (e.g., the similarity of activation during face blocks vs. activation during house blocks) [56]. We use correlation coefficients to measure similarity. We then average all the within-condition correlations for a given task and subtract the average of all the between-condition correlations in that same task and use that as our measure of neural distinctiveness.

#### Fixel-based DWI Analysis

To analyze diffusion weighted imaging data, we are using fixel based analyses (FBA) which computes estimates of fiber density (FD), fiber bundle cross-section (FC), and fiber density and cross section (FDC) throughout each participant’s white matter mask. Calculations of FD, FC, and FDC are outlined in <u>Dhollander et al.</u> [57]

### Statistical Analysis

#### Aim 1: Longitudinal Analysis

Baselines and trajectories of change in neural distinctiveness, GABA, and behavior will be mapped across age using multilevel growth curve models. These models are referred to as multilevel (mixed-effects, hierarchical) models because they include both fixed and random effects and nest repeated observations within people in order to capture individual trajectories of change with age. The fixed effects are parameter estimates that apply to the entire sample and do not vary across individuals. In our models, we will estimate two fixed effects: the group intercept and the group slope. The group intercept is an estimate of the average starting point of the trajectory. Because all of our participants will be at least 65 years old, our group intercepts will be estimates of the average value of a variable (e.g., GABA, neural distinctiveness, a behavioral measure) at age 65. The group slopes estimate the average rate of change in a variable with age. These two fixed effects capture the typical trajectory of a variable with increasing age in the entire sample. In the current project, we will test for longitudinal declines in GABA, in neural distinctiveness, and in behavior simply by testing whether the fixed effect of slope for that variable is significantly negative. In contrast to the fixed effects, the random effects represent between-subject variability around the fixed effects. A small random effect of intercept means that individuals in the sample have similar intercepts while a large random effect means greater variability in intercepts. Likewise, a small random effect of slope means similar rates of change with age across subjects while a large random effect means high variability in slopes.

#### Aim 2: Comparing MCI patients to Age-Matched Controls

We will use a general linear model to investigate whether our measures (e.g., neural distinctiveness, GABA, behavioral scores, DWI measures) are significantly different in the MCI patients than in age-matched control participants. We will analyze the participant group (controls vs. MCI patients) as the categorical variable of interest, but will also include age and sex as additional covariates (as well as regional grey matter volume if appropriate).

#### Aim 2: PET Data Analysis

We will use FreeSurfer and MRS voxel placements in each region of interest (ROI; 7 total), to analyze amyloid and tau concentrations in those ROI’s. In addition, we will use SPM to coregister anatomical masks for each participant and their PET scan. We will compare neural distinctiveness values in each ROI, per participant, with their amyloid and tau concentrations.

## Discussion

This study is innovative in at least five important ways. First, we will collect fMRI, MRS, and behavioral measures in the same participants, allowing us to directly investigate the relationship between neural distinctiveness (assessed using fMRI), GABA levels (assessed using MRS), and age-related behavioral declines (assessed using psychophysical and assessment techniques).

Second, we will collect longitudinal data across a period of almost 10 years, with 3 time points in many participants. By employing a longitudinal design, we will be able to examine within-participant trajectories of change with aging rather than just cross-sectional age differences. In addition, our results will not be confounded by cohort effects, and participants will serve as their own control, which maximizes power.

Third, the longitudinal data that we collect will allow us to examine directional relationships between our measures of interest. Rather than simply calculating zero-order correlations between different measures acquired at a single datapoint, the dataset we have outlined will allow us to explore more mechanistic directional questions, such as whether GABA levels at age 65 predict subsequent declines in neural distinctiveness, or whether measures of neural distinctiveness at age 65 predict subsequent changes in behavioral performance.

Fourth, we will use state of the art multilevel growth curve models that gracefully handle missing data, attrition, and different time delays, all of which we likely will experience during data collection. Whereas traditional approaches such as repeated measures ANOVA and difference scores require every participant to have data from every time point, growth curve models exploit full information maximum likelihood (FIML) to create probable values for missing data, rather than deleting missing participants or requiring imputation. The use of these models means that we will be able to include every measure from every participant, even if that participant had fewer time points than others, or left the study early. Use of such models will significantly increase our statistical power.

Fifth, we will explore GABA-based neural dedifferentiation in the context of Alzheimer’s pathology, by recruiting MCI patients who have already undergone PET imaging sessions to assess amyloid beta and tau burdens. By testing whether individual differences in GABA are associated with individual differences in distinctiveness, and whether distinctiveness mediates the relationship between Alzheimer’s pathology and behavior, explains additional behavioral variance over and above the neuropathology, or both, we hope to further understand the neural mechanisms underlying behavioral impairments in Alzheimer’s disease.

Of course, there are limitations to the study. One limitation is that the first datapoint in our longitudinal dataset is acquired from participants recruited at age 65 or older and that it is possible changes in neural distinctiveness and GABA begin earlier than this. Indeed, an investigation of the trajectory of cortical GABA demonstrates a decline which begins in middle age, at approximately 40 years [58].

Another limitation is related to learning effects. In order to estimate the trajectories of behavioral change, we will administer the same battery of behavioral tasks at each time point. However, participants could plausibly exhibit practice effects. Since the time between administrations of this battery is quite long (typically 3 years or more) we anticipate that practice effects are likely to be small. In addition, analyses investigating whether GABA or neural distinctiveness predict subsequent behavioral change are about individual differences in behavioral change, which are less likely to be systematically influenced by practice effects.

Despite the few limitations, the MiND study’s protocol is novel and innovative in aging research by collecting longitudinal data and studying Alzheimer’s pathology alongside healthy older adults with a focus on neural distinctiveness. By utilizing these methods, we plan to uncover more about the role of neural dedifferentiation and GABA in age-related cognitive decline.

## Data Availability

Data sharing is not applicable to this article as no datasets were generated or analyzed during the current study.

## Abbreviations

MiND: Michigan Neural Distinctiveness Study
fMRI: Functional magnetic resonance imaging
MRS: Magnetic resonance imaging
GABA: gamma-aminobutyric acid
MCI: Mildly Cognitively Impaired
DAT: Dementia Alzheimer’s Type
PET: Positron Emission Tomography
NACC UDS3: National Alzheimer’s Coordinating Center Uniform Data Set Version 3
NIA: National Institute on Aging
NIH: National Institute of Health
P30: Center Core Grants
MADRC: Michigan Alzheimer’s Disease Research Center
BMH: Board Certified Clinical Neuropsychologist
GDS: Geriatric Depression Scale
MoCA: Montreal Cognitive Assessment
MINT: Multilingual Naming Test
fTORT: Functional Tactile Object Recognition
WAIS: Wechsler Adult Intelligence Scale
RMBT: Rivermead Behavioral Test
VPA: Verbal Paired Associates
DWI: Diffusion Weighted Imaging
VOI: Voxel of Interest
GLM: Generalized Linear Model
FSFAST: FreeSurfer Functional Analysis Stream
FWHM: Full Width Half Maximum
BOLD: Blood Oxygen Level Dependent
FD: Framewise Displacement
OLS: Ordinary Least Squares
ROI: Region of Interest
ICA: Independent Component Analysis
FOD: Fiber Orientation Distributions
IU: Institutional Units
CSF: Cerebrospinal Fluid
MVPA: Multi-voxel Pattern Analysis
FBA: Fixel Based Analysis
FDC: Fiber Density and Cross Section
FIML: Full Information Maximum Likelihood

## Declarations

### Ethics approval and consent to participate

In accordance with the University of Michigan’s Human Research Protection Program (HRPP), the MiND project is approved by the University of Michigan Medical School Institutional Review Board (IRBMED; Study ID HUM00199054). Participants provide verbal consent during screening interview and written consent at the beginning of Session 1.

### Consent for Publication

Not Applicable

### Author’s Contributions

TP is the PI of the study and received the grant funding the project. EK and MS were the main authors of the manuscript. EK, NR, and KW were major contributors to the coordination of the project and data collection. QZ, KW, and MZ reviewed sections of the paper for accuracy and details. MS, AMB, BMH, SFT, TP were the main contributors to task design and data analysis. All authors read, reviewed, and approved the final manuscript.

## Acknowledgements

Not Applicable

